# Young Plasma Infusions Significantly Improve Clinical Symptoms and UPDRS Scores in Patients with Parkinson’s Disease

**DOI:** 10.64898/2026.05.12.26353041

**Authors:** Dian J Ginsberg, Carl Scheffey

## Abstract

In both preclinical and clinical studies, transfusions of plasma from young individuals have been reported to ameliorate aspects of neurodegeneration. This study was designed as a preliminary test of the hypothesis that plasma transfusions from young donors might benefit Parkinson’s patients. 19 patients were allocated to receive either 2-liters of plasma from young donors, in two doses spaced two days apart, or two doses of placebo. For the next 24 weeks, this double-blind study evaluated changes on a modified MDS-UPDRS scale, along with blood tests and other observations. Adverse events possibly related to transfusion were mild rise in blood pressure and urticaria. A t-test on the changes in the sum of UPDRS subscales 1-3 showed that the plasma patients did better than the placebo patients (p = 0.03*). For patients given yFFP® (young Fresh Frozen Plasma), the estimated decrease in the sum of scales 1-3 was 7.1 (95% conf. interval 4.3 to 9.9). Our results give a preliminary indication that young plasma transfusions reduce Parkinson’s symptoms and have a place in treatment of these patients. (NCT 04202757).

## 1. Introduction

Medicine has been attempting to find a cure for Parkinson’s Disease (PD) since James Parkinson described the ‘Shaking Palsy’ in 1817[1]. Treatment to this date has been targeted medication ‘at’ the neuronal dysfunction that enables the patient to function as best as possible while the disease progresses. Gene therapy is difficult as many limitations hamper the successful development of these candidates, including the need for difficult surgical guided placement, as well as the lack of biomarkers available for diagnosis and therapy management[2,3].

A common feature of PD and dementias is chronic neuroinflammation, mitochondrial dysfunction and oxidative stress[4–6]. Neuromelanin synthesis is a protective antioxidant mechanism in the brain to remove potentially toxic oxidized byproducts in the substantia nigra. The protective mechanism can be overburdened as toxic load becomes unsustainable for the cell. Alpha-synuclein and Lewy bodies build up, destroying its proteostasis, mitochondrial function and overall ability to survive. Eventually enough of the dopaminergic neurons in the substantia nigra will die and PD symptoms will develop.

Preclinical and clinical research have recently accumulated evidence that the plasma of younger individuals appears to contain rejuvenating properties that can have an impact on degenerative disease. In the present paper, we refer to the plasma of younger individuals as “Young Plasma.” In clinical studies, we use the acronym yFFP (young Fresh Frozen Plasma). The evidence raises the question: can neurodegenerative diseases be reversed? Molecular pathways lost in neurodegenerative disease might be reinstated by “the transfer of youthful factors into relevant neural niches” [7]. In the mouse hippocampus, Young Plasma has been reported to reverse a variety of age-related changes[8]. Similar improvements have been seen in other rodent trials [9–13]. Clinical human studies have reported related findings with yFFP or plasma exchange[14–17].

The safety of transfusions with plasma from young donors is already established. The American Red Cross estimates 10,000 units of plasma are used daily in the United States[18]. An estimated 19.8% of blood donations came from the age group 16-24 years, and transfusions have had an excellent safety record[19,20].

The primary objective of our study was to assess effectiveness of infusions of two liters of yFFP on Parkinson’s patients. The primary objective of our study was to assess effectiveness of infusions of two liters of yFFP on Parkinson’s patients. The exploratory objectives were clinical improvements using: UPDRS, the Stanford Presenteeism Score (SPS-6)[21], and laboratory parameters.

## 2. Methods

### 2.1. Enrollment and treatment allocation

The target enrollment for the study was 20 patients. Inclusion criteria were: mild to moderate Parkinson’s Disease (less than 3 on Hoehn-Yahr scale), with the ability to do a self-evaluation or have a caregiver to perform the evaluation. Patients with certain severe comorbidities were excluded. The study’s protocol and informed consent form were evaluated and approved by the Institute of Cellular and Regenerative Medicine (IRCM) in Santa Monica, Ca. IRB organization number IORG0007913. Patients to be enrolled had all their questions answered and signed an informed consent to participate in the study.

The treatment under study was two transfusions of yFFP given on each of two days, with one intervening day between the first and second transfusions. The volume of each transfusion was

12.5 ml-per-kg^-1^. Donors were matched with patients by blood type and sex. Plasma was slowly infused in order to minimize transfusion-associated circulatory overload (TACO). Patients allocated to placebo received the same volume of fluid on the same schedule, but the fluid was isotonic NaCl with 0.1% pyridoxine (vitamin B6). This enabled the fluid to be a yellow color to even further ensure that even with the mask bag cover the patient would be blinded.

The patients and all persons who had direct contact with the patients were blinded as to the treatment allocation. That includes the neurologists who did the outcome measure evaluations.

When possible, alternating consecutive patients were allocated to receive yFFP or placebo. As yFFP can be in short supply and blood type matching is critical, it was understood that allocation to yFFP might depend upon the blood type available.

### 2.2. Plasma Collection

FDA approved [21CFR640.30] Plasma was collected from sex-identified individuals 18-25 years of age (yFFP®), by Spectrum Plasma. Apheresis machines were used for all collections, which were immediately processed and then held frozen at −30°C. In addition to the twelve standard plasma tests required by FDA regulation (21CFR610.40), Spectrum Plasma’s standard operating procedure is to perform five more tests to ensure safety: all candidates are pretested before becoming eligible donors; mean corpuscular volume, hemoglobin and hematocrit are tested; a serum protein electrophoresis test is performed before a donation is made to confirm the donor has acceptable total protein levels; all plasma collected from female donors is tested for the presence of HLA antibodies; to avoid latent infections, yFFP is held for a minimum of thirty days, within which the donor is required to make another donation that passed all tests, before the first donation is released. All shipments are frozen, temperature monitored and then warmed at the time and point of use.

### 2.3. Data Collection

The explored outcomes were documented at baseline then at 4, 12, and 24 weeks. Evaluations were done via a modified form of MDS-UPDRS, the Stanford Presenteeism Scale (SPS-6), and laboratory parameters. Additionally, each patient was also given a blank sheet of paper and the option to journal their overall feelings and any changes they personally wanted to share.

The UPDRS scale used was modified for subscale 3. The modified form used in the UPDRS subscale 3 recorded only one score for certain groups of questions. For example, only one score was recorded for the first five questions of the subscale, all of which are on the subject of resting tremor. The score recorded was the maximum for the answers to the five questions. For bradykinesia questions with two bilateral scores, only the maximum of the two scores was recorded. Accordingly, the maximum for subscale 3 was 56. The minimum clinically important difference for the UPDRS scale used was not the same as for the full MDS-UPDRS.

### 2.4. Statistical Analysis

In advance of computing any statistics, the authors agreed that any p-value less than 0.1 (with two-sided alternative) would be considered a preliminary result worth further exploration. We reserve the phrase “statistically significant” to mean p < 0.05.

Statistics in this study were based on comparing the outcome measures collected at baseline with those at 24 weeks. The change in scores was evaluated for the two groups of patients with each treatment. Additionally, the populations of changes in the two groups were compared with each other.

For both of those kinds of comparisons, the null hypothesis was of zero difference between two group means. For each comparison, we report two-sided p-values for a permutation test[21]. Monte Carlo computations used 10^8^ permutations. For each permutation, the test statistic was the difference between means in the two subpopulations of the pooled data. We also report results of t-tests (with the equal variance assumption), which are parametric, along with parametric estimates for the differences in means between the compared groups. The t-test is familiar to more readers, and affords comparison between the present study and the many studies in the literature that have used t-tests to interpret changes in the UPDRS and SPS scales. Strictly speaking, the assumptions of the t-test are not satisfied for these discrete data (see Fig. 2). It is, nevertheless, a useful tool. The permutation test is nonparametric, and makes weaker assumptions about the origins of the data. Each population was checked for data characteristics such as extreme skewness that render permutation tests inappropriate[22].

Statistical computations used the R statistical language[23], with the “coin” add-on package[24].

## 3. Results

### 3.1 Enrollment

Fig. 1 shows a CONSORT-style patient flow diagram. Of the 22 patients enrolled, three withdrew for personal reasons prior to study initiation. Nine patients were allocated to receive yFFP, and ten patients were allocated to placebo.

**Figure 1.**
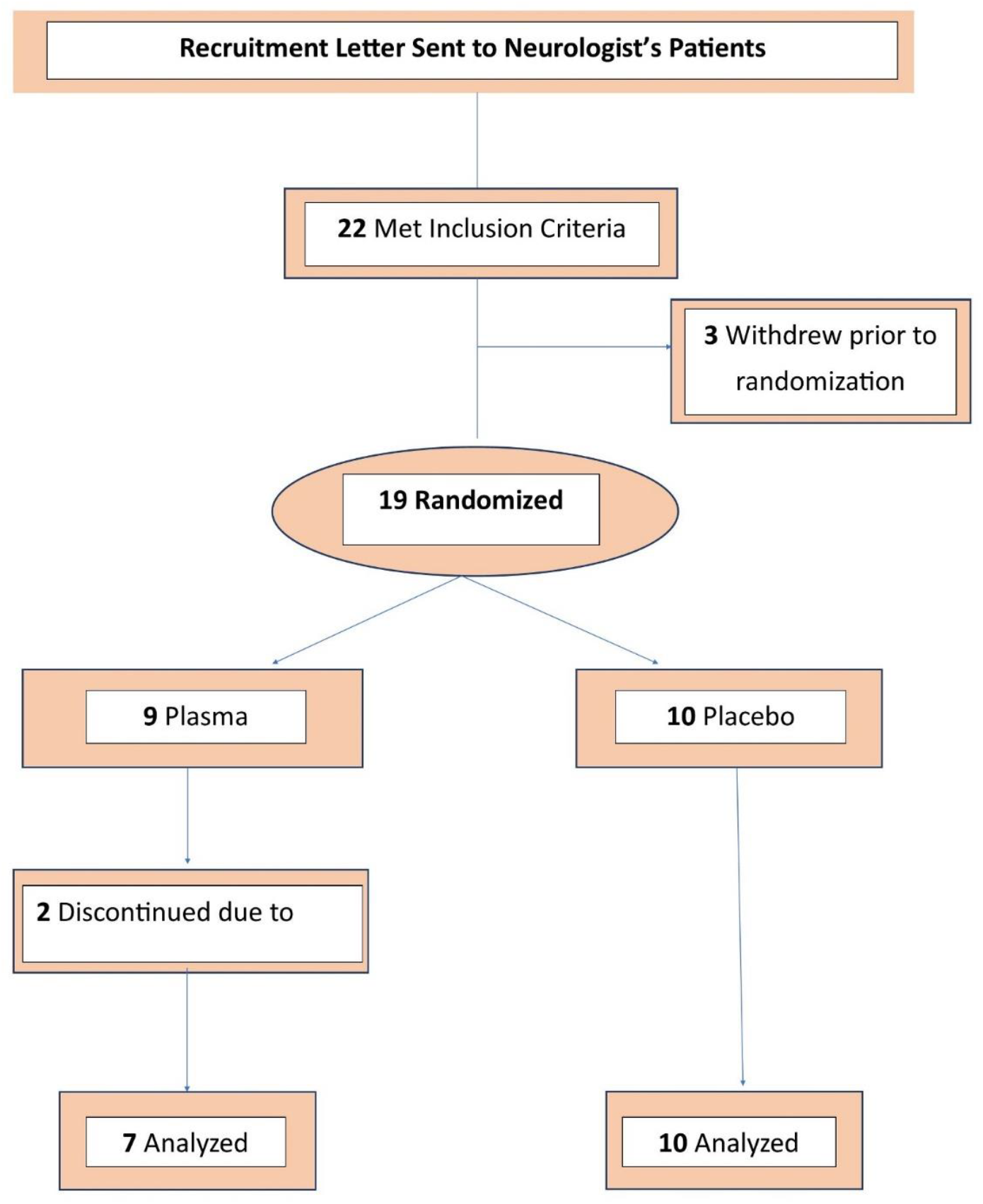
Diagram of Patient Enrollment and Analysis.

**Fig. 2.**
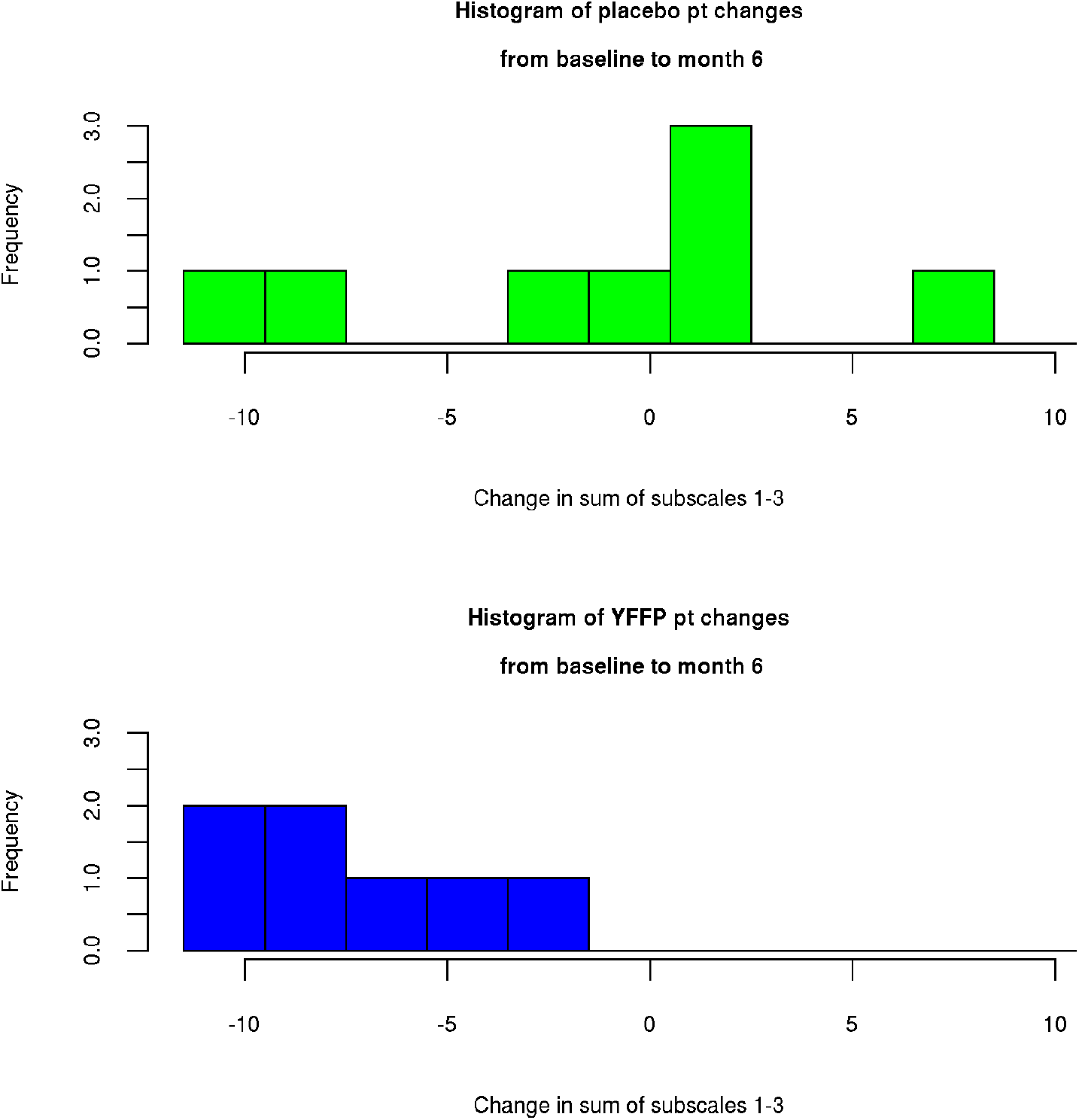
Histograms of changes in sum of UPDRS subscales.

### 3.2. Baseline Patient Characteristics

Tables 1 and 2 compare the treatment groups with respect to certain demographics and baseline patient characteristics. Most of the patient samples were males over the age of 60 years. There were not dramatic differences seen between the allocation groups.

**Table 1.** Baseline patient characteristics (part 1).

|  | % male | %smoke | % any alcohol | taking<br>carbidopa |
| --- | --- | --- | --- | --- |
| yFFP | 86% | 0% | 56% | 57% |
| placebo | 80% | 10% | 50% | 25% |

**Table 2.** Baseline patient characteristics (part 2).

|  | treatment | Q1 | median | Q3 |
| --- | --- | --- | --- | --- |
| Weight (kg) | yFFP | 48 | 82 | 114 |
|  | placebo | 63 | 80 | 93 |
| Age | yFFP | 61 | 68 | 70 |
|  | placebo | 60 | 70 | 72 |
| sum UPDRS<br>scales 1-3 | yFFP | 16 | 30 | 32 |
|  | placebo | 18 | 28 | 35 |
| Hohn-Yahr | yFFP | 1 | 1.5 | 2.25 |
|  | placebo | 1.125 | 1.5 | 1.5 |
| SPS | yFFP | 12.75 | 15.5 | 17.5 |
|  | placebo | 15 | 16 | 17 |

### 3.3 Changes on individual scales

For each scale, the patient scores were compared between baseline and 24 weeks. The results are shown in Table 3. For patients treated with yFFP, there were statistically significant decreases in UPDRS subscales 1 and 2.

**Table 3.** Statistics comparing baseline values with 24-week values, within each treatment group.

| scale | treatment | permutation test<br>p-value | paired t-test<br>p-value | parametric estimate,<br>end minus baseline<br>(95% conf. int.) |
| --- | --- | --- | --- | --- |
| sum | yFFP | 0.0006*** | 0.0008*** | -7.1 (-9.9, -4.3) |
| UPDRS<br>subscales 1-3 |  |  |  |  |
|  | placebo | 0.829 | 0.744 | 1.0 (-5.4, 7.3) |
| sum | yFFP | 0.0005*** | 0.005** | -5.3 (-8.3, -2.3) |
| UPDRS<br>subscales 2-3 |  |  |  |  |
|  | placebo | 0.681 | 0.619 | 1.4 (-4.8, 7.6) |
| UPDRS | yFFP | 0.021* | 0.034* | -1.3 (-3.4, -0.2) |
| subscale 1 |  |  |  |  |
|  | placebo | 0.211 | 0.193 | -0.5 (-1.2, 0.3) |
| UPDRS | yFFP | 0.0006*** | 0.004** | -3.6 (-5.5, -0.2) |
| subscale 2 |  |  |  |  |
|  | placebo | 0.783 | 0.692 | -0.5 (-3.3, 2.3) |
| UPDRS | yFFP | 0.364 | 0.370 | -1.7 (-6.0, 2.6) |
| subscale 3 |  |  |  |  |
|  | placebo | 0.351 | 0.285 | 1.9 (-1.9, 5.7) |
| SPS |  |  |  |  |
|  | yFFP | 0.061<sup>a</sup> | 0.048* | -3.6 (-7.3, -0.1) |
|  | placebo | 0.559 | 0.397 | -0.5 (-1.4, 0.4) |

### 3.4. Comparing UPDRS and SPS Changes

Table 4 shows a more direct comparison of yFFP with placebo. For each patient, the score on that scale at baseline was subtracted from the score at 24 weeks. That difference was the change in the measure during the study. The population of those differences for yFFP patients was statistically compared with the population of those differences for placebo patients. The comparison was statistically significant for the sum of UPDRS subscales 1-3, and for the SPS scale.

**Table 4.** Comparing treatment groups for size of change from baseline to 24 weeks.

| scale | permutation test<br>p-value | t-test<br>p-value |
| --- | --- | --- |
| sum UPDRS scales 1-3 | 0.025* | 0.039* |
| sum UPDRS scales 2-3 | 0.058<sup>a</sup> | 0.71 |
| UPDRS scale 1 | 0.082<sup>a</sup> | 0.062<sup>a</sup> |
| UPDRS scale 2 | 0.081<sup>a</sup> | 0.076<sup>a</sup> |
| UPDRS scale 3 | 0.184 | 0.167 |
| SPS | 0.022* | 0.017* |

Figures 2 and 3 show histograms of the some of the differences that were used for the statistical tests of Table 4. Figure 2 shows the differences for the sum of all three of the UPDRS subscales 1-3. Fig. 3 shows the differences for the SPS-6 scale. In each pair of histograms, the top half of the figure shows the distribution of placebo changes from baseline to 24 weeks, and the bottom histogram shows the distribution of yFFP changes. In Fig. 2, more of the yFFP patients showed improvement (negative numbers) for the UPDRS at 24 weeks, minus baseline. Fig. 3 shows a similar comparison for the changes in SPS-6 scores.

**Fig. 3.**
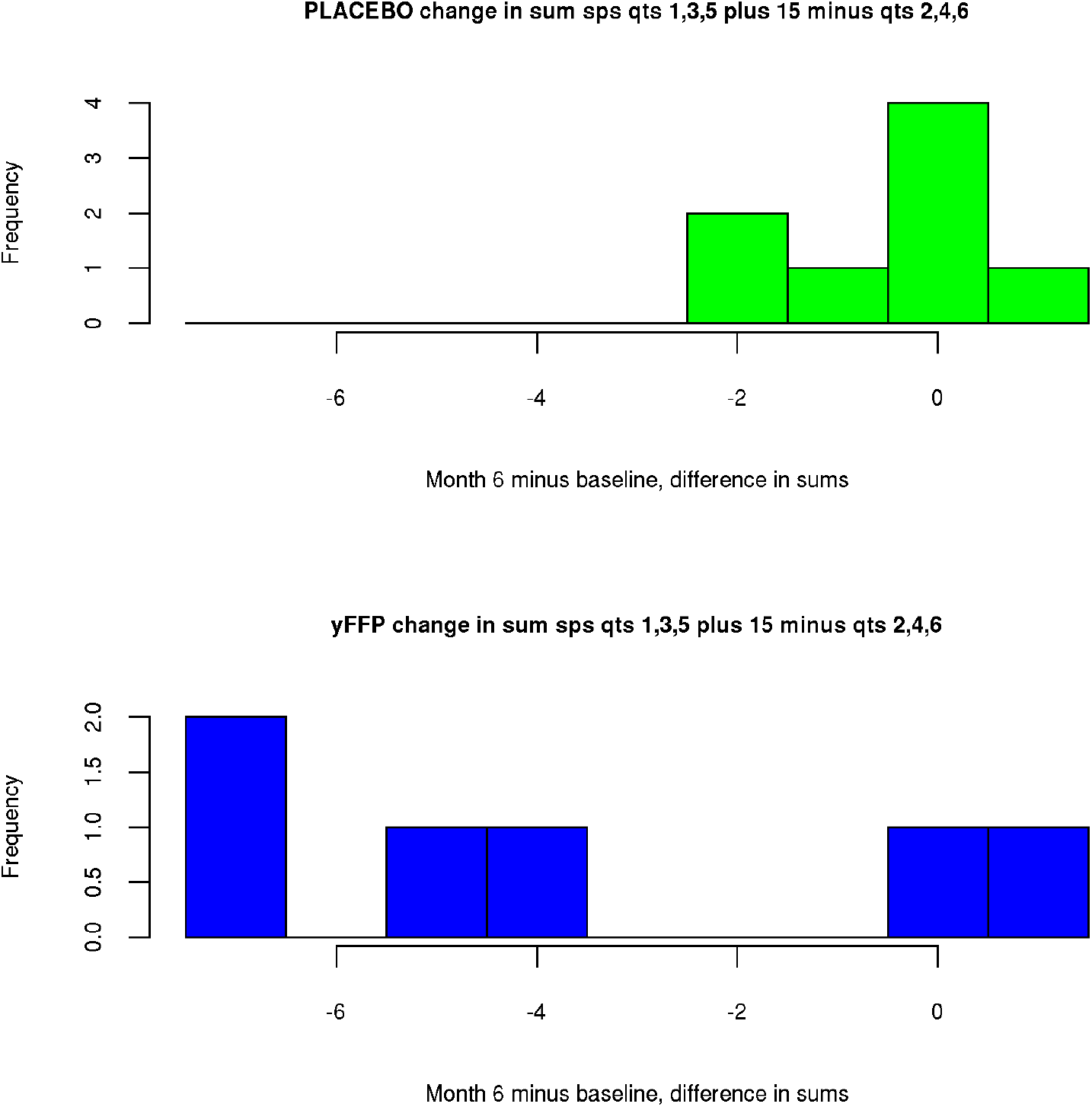
SPS-6 score change histograms.

### 3.5. Safety

There were 5 total mild adverse events in the plasma group and none in the placebo group. Two of the patients, one with hives and one with a rise in blood pressure, chose with the nurse not to complete the infusions and subsequently withdrew from the study. Of the other 3 patients, one patient who was already on hypertensive medications had a small rise in blood pressure (baseline was 132/77 mm Hg (17.6/10.3 kPa) and max was 165/87 mm Hg (22.0/11.6 kPa). The blood pressure normalized before the completion of the infusion. Two patients experienced some itching but neither showed signs of urticaria and all symptoms resolved with IV diphenhydramine and oral fexofenadine. None of the patients had any deleterious changes in their laboratory values.

## 4. Discussion

This study presents evidence that the administration of young Fresh Frozen Plasma significantly improves multiple outcome measures during the 24 weeks after administration. The comparisons of Table 4 show statistically significant results that patients assigned to yFFP improved more than patients assigned to placebo, with respect to the sum of UPDRS subscales 1-3 and also SPS-6.

The most common risks associated with FFP include: transfusion related acute lung injury, circulatory overload, anaphylactic reactions, transmission of infections and hemolytic transfusion reactions[25]. Because precautions were taken to avoid those risks, we believe that most appropriate statistical analysis would assume a priori that yFFP could not worsen the outcome measures, in the general pool of patients used for enrollment. Under that assumption, one-sided rather than two-sided tests would be appropriate and p-values would be cut in half. We note in tables 3 and 4 the tests that then become statistically significant in the sense of p < 0.05.

As the dopamine loss within the central nervous system creates lack of both muscle use and imbalanced forces over time, peripheral dysfunction in the muscle will occur. Therefore, the motor changes may require more time and muscle rehabilitation to show a significant improvement[26,27]. We hypothesize that the plasma crossing the blood brain barrier partially reversed the inflammatory process of the neurodegenerative disease. It may be that yFFP replaces as well as modulates production of these factors in the brain such as brain derived neurotrophic factor, antioxidant support, oxytocin, thrombospondin-4 and SPARC-like protein 1 as well as upregulating enzymes such as enzyme ten eleven translocation methylcytosine dioxygenase 2 (Tet2), known to support neurogenesis[28–31]. Many of these components have a very short half-life so consideration that supplying smaller aliquots of yFFP weekly perhaps will be a better future option. As apheresis and the removal of senescent and inflammatory cells has been reported to give very promising results[15,32], combining this removal process with the addition of young factors present only in yFFP may prove to be optimal therapy to address both the improvement of disease states as well as provide a mechanism for healthy aging. We refer to this combination approach as ‘Rectangularization of Aging.’

The young Fresh Frozen Plasma (yFFP) used in this study was collected by methods intended to safeguard the factors described in the above paragraph. In general, typical blood bank plasma is collected from donors of all ages without sex-segmentation, is often kept for up to eight hours as whole blood and in some collections, treated with methylene blue and UV light. It has been shown that those legal, but inferior methods of collection can incur as much as a 58% reduction in important factors and proteins[33,34]. It has also been shown that there are significant differences in the plasma proteome between sexes and ages[35].

Laboratory parameters did not show that the patients had any harmful effects from the plasma. While our study did have 2 patients show a mild reaction requiring discontinuation of the infusion, no severe reactions were recorded.

This preliminary study has limitations. The sample was small. The plasma/placebo treatment allocation was largely done by alternation between consecutive patients, rather than randomization. UPDRS evaluations were not done at a consistent time of day and were scheduled without regard for the medication schedules of the individuals. Patients were recruited via an email sent from two large neurology practices, and the volunteers who qualified became the sample.

In spite of the study limitations, the results support the hypothesis that yFFP has benefit to Parkinson’s patients. Further studies with larger groups of patients and comparing therapies need to be explored.

## Data Availability

All data produced in the present work are contained in the manuscript

https://www.youngplasmastudy.com

## Acknowledgements

Patient evaluations were performed by the neurologists: Igor Cherches, MD and Eddie Patton Jr, MD. This work was supported by a grant from the Carolina Longevity Institute. Spectrum Plasma donated the yFFP and supported preparation of the manuscript. The sponsors and participating physicians all contributed to the study design. No sponsor analyzed or contributed to the interpretation of any data.

## Declaration of Interest

Dian J. Ginsberg, MD is a shareholder in Spectrum Plasma, Inc.

Carl Scheffey, PhD received consulting fees from Spectrum Plasma, Inc.

The acronym yFFP is a registered trademark of Spectrum Plasma, Inc.

## Authors contributions

**Dian J. Ginsberg, MD:** Supervision, Funding acquisition, Project administration, Conceptualization, Methodology, Investigation, Data curation, Visualization, Writing-original and review.

**Carl Scheffey:** Formal analysis, Software, Methodology, Data curation, Visualization, Writing--original and review.

